# Investigating gene-diet interactions impacting the association between macronutrient intake and glycemic traits

**DOI:** 10.1101/2022.07.26.22278077

**Authors:** Kenneth E. Westerman, Maura E. Walker, Sheila M. Gaynor, Jennifer Wessel, Daniel DiCorpo, Jiantao Ma, Alvaro Alonso, Stella Aslibekyan, Abigail S. Baldridge, Alain G. Bertoni, Mary L. Biggs, Jennifer A. Brody, Yii-Der Ida Chen, Joseé Dupuis, Mark O. Goodarzi, Xiuqing Guo, Natalie R. Hasbani, Adam Heath, Bertha Hidalgo, Marguerite R. Irvin, W. Craig Johnson, Rita R. Kalyani, Leslie Lange, Rozenn N. Lemaitre, Ching-Ti Liu, Simin Liu, Jee-Young Moon, Rami Nassir, James S. Pankow, Mary Pettinger, Laura Raffield, Laura J. Rasmussen-Torvik, Elizabeth Selvin, Mackenzie K. Senn, Aladdin H. Shadyab, Albert V. Smith, Nicholas L. Smith, Lyn Steffen, Sameera Talegakwar, Kent D. Taylor, Paul S. de Vries, James G. Wilson, Alexis C. Wood, Lisa R. Yanek, Jie Yao, Yinan Zheng, Eric Boerwinkle, Alanna C. Morrison, Miriam Fornage, Tracy P. Russell, Bruce M. Psaty, Daniel Levy, Nancy L. Head-Costa, Vasan S. Ramachandran, Rasika A. Mathias, Donna K. Arnett, Robert Kaplan, Kari E. North, Adolfo Correa, April Carson, Jerome Rotter, Stephen S. Rich, JoAnn E. Manson, Alexander P. Reiner, Charles Kooperberg, Jose C. Florez, James B. Meigs, Jordi Merino, Deirdre K. Tobias, Han Chen, Alisa K. Manning

**Author notes:** co-corresponding authors **Corresponding Authors:** Kenneth E. Westerman, Alisa K. Manning.

## Abstract

**Background:** Heterogeneity in the long-term metabolic response to dietary macronutrient composition can be partially explained by genetic factors. However, few studies have demonstrated reproducible gene-diet interactions (GDIs), likely due in part to measurement error in dietary intake estimation as well as insufficient capture of rare genetic variation. Discovery analyses in ancestry-diverse cohorts that include rare genetic variants from whole-genome sequencing (WGS) could help identify genetic variants modifying the effects of dietary macronutrient composition on glycemic phenotypes.

**Objective:** We aimed to identify macronutrient GDIs across the genetic frequency spectrum associated with continuous glycemic traits in genetically and culturally diverse cohorts.

**Methods:** We analyzed N=33,187 diabetes-free participants from 10 cohorts in the NHLBI Trans-Omics for Precision Medicine (TOPMed) program with WGS, self-reported diet, and glycemic traits (fasting glucose [FG], insulin [FI], and hemoglobin A1c [HbA1c]). We fit multivariable-adjusted linear mixed models for the main effect of diet, modeled as an isocaloric substitution of carbohydrate for fat, and for its interactions with genetic variants genome-wide. Tests were performed for both common variants and gene-based rare variant sets in each cohort followed by a combined cohort meta-analysis.

**Results:** In main effect models, participants consuming more calories from carbohydrate at the expense of fat had modestly lower glycemic trait values (β per 250 kcal substitution for FG: −0.030 mmol/L, *p*=2.7×10^−6^; lnFI: −0.008 log(pmol/L), *p*=0.17; HbA1c: −0.013 %, *p*=0.025). In GDI analyses, a common African ancestry-enriched variant (rs79762542; 78 kb upstream of the *FRAS1* gene) reached study-wide significance (*p* = 1.14×10^−8^) indicating a higher HbA1c with greater proportion of calories from carbohydrate vs. fat among minor allele carriers only. This interaction was replicated in the UK Biobank cohort. Simulations revealed that there is (1) a substantial impact of measurement error on statistical power for GDI discovery at these sample sizes, especially for rare genetic variants, and (2) over 150,000 samples may be necessary to identify similar macronutrient GDIs under realistic assumptions about effect size and measurement error.

**Conclusions:** Our analysis identified a potential genetic interaction modifying the dietary macronutrient-HbA1c association while highlighting the importance of precise exposure measurement and significantly increased sample size to identify additional similar effects.

## INTRODUCTION

Diet is an established modifiable factor associated with risk of type 2 diabetes (T2D) and related cardiometabolic diseases(1). However, evidence is mixed regarding the ideal dietary macronutrient composition for risk reduction. Dietary interventions with differing proportions of energy from carbohydrates versus fat have shown varied efficacy for T2D risk reduction and substantial between-person heterogeneity in effects on cardiometabolic risk factors(2–4). Further, acute glycemic responses to meals with specific macronutrient composition are reproducible within individuals(5,6). Genetically different mouse strains have varying sensitivity of glycemic biomarkers to a high-fat diet(7) and to human-relevant dietary patterns(8). Retrospective analysis of human trials manipulating macronutrient intake have found genetic modifiers of glycemic response(9). Taken together, such studies suggest that genetics could be a key contributor to variability in the association between dietary macronutrient composition and glycemic health.

Gene-diet interaction (GDI) studies aim to identify genetic variants that modify the association between dietary behaviors and health. Furthermore, GDI studies support differential associations of dietary factors with glycemic traits according to genotypes, using both hypothesis-driven(10) and hypothesis-free(11,12) strategies. However, in general, discovery and replication of gene-diet interactions with T2D risk has been poor, possibly due to measurement error in assessing habitual diet, low statistical power for interaction analysis, and biological and behavioral heterogeneity across populations(13). Additionally, to date there has been little exploration of GDIs involving rare genetic variants, which affect a smaller proportion of the population but may have larger effect sizes(14).

Our primary aim was to discover novel putative genetic modifiers for the association between dietary macronutrient composition and glycemic traits. To this end, we performed a GDI analysis using common and rare genetic variants in over 30,000 individuals with diverse ancestral backgrounds from the NHLBI Trans-Omics for Precision Medicine (TOPMed) program. We focused on modeling a dietary carbohydrate-fat exchange, which can be reasonably assessed via self-reported diet questionnaires and can be straightforwardly modified in the context of a healthful diet. Furthermore, the use of whole-genome sequencing (WGS) permitted the analysis of rare variants using set-based association tests. As a secondary aim, we sought to inform subsequent GDI research by exploring the impact of dietary exposure measurement error on statistical power in the context of realistic effect size estimates.

## RESULTS

We analyzed data from 33,178 individuals without diabetes (based on FG, HbA1c, or medication use) from 10 TOPMed program cohorts. Participants had diverse cohort-reported race/ethnicities including: African American (N = 6,158), American Indian (N = 35), Asian (N = 124), white (N = 19,721), and Hispanic/Latino (N = 7,114). Dietary carbohydrate and fat as a percentage of total energy intake on average were 50.5% (standard deviation = 8.5%) and 32.2% (6.9%), respectively, in the full pooled sample, estimated using validated food frequency questionnaires (FFQ) or 24-hour dietary recalls (24HR). Cohort-specific carbohydrate intake estimates (as percent of total energy [% kcal]), glycemic trait values (fasting glucose [FG], fasting insulin [FI; or log-transformed, lnFI], and hemoglobin A1c [HbA1c]), and additional population characteristics are presented in Supp. Table S1 and Supp. Fig. S1.

We first modeled the main association of macronutrient compositions with each of the glycemic traits. By adjusting for total energy and energy from protein, resulting regression estimates for carbohydrate represented a macronutrient exchange (increased 250kcals from carbohydrate replacing an equivalent 250kcals from fat; see Methods). Meta-analysis of the individual cohorts indicated that a higher proportion of kcal from carbohydrate at the expense of fat was associated with lower FG (−0.030 mmol/L / 250kcals; *p* = 2.2×10^−6^), lnFI (−0.008 log(pmol/L) / 250kcals; *p* = 0.15), and HbA1c (−0.012 %HbA1c / 250kcals; *p* = 0.029). Forest plots of these results are shown in Supp. Fig. S2.

### Common variant gene-diet interactions

We sought to identify macronutrient GDIs with the maximal sample available in TOPMed program cohorts in order to provide a baseline for discovery and evaluate our assumptions about expected effect sizes. Common variants (MAF > 1%) were analyzed in a primary, single-variant analysis of gene-carbohydrate interactions, with the same regression adjustments as above. This GDI analysis produces interaction estimates for the difference in the macronutrient-glycemic trait association per alternate allele at the variant of interest. After genome-wide, cohort-specific analysis and cross-cohort meta-analysis, one variant reached a study-wide significance threshold of 1.67×10^−8^ (5×10^−8^ / 3 glycemic traits). Two additional variants passed a standard genome-wide threshold of 5×10^−8^ (Table 1). We note that this threshold is liberal given the greater testing burden involved in the analysis of multiple ancestry groups(15). Of these three, none had evidence of a genetic main effect on the associated trait. Results are visualized in Supp. Fig. S3 for all variants and shown in Supp. Table S3 for variants with interaction *p* < 10^−5^.

**Table 1:**
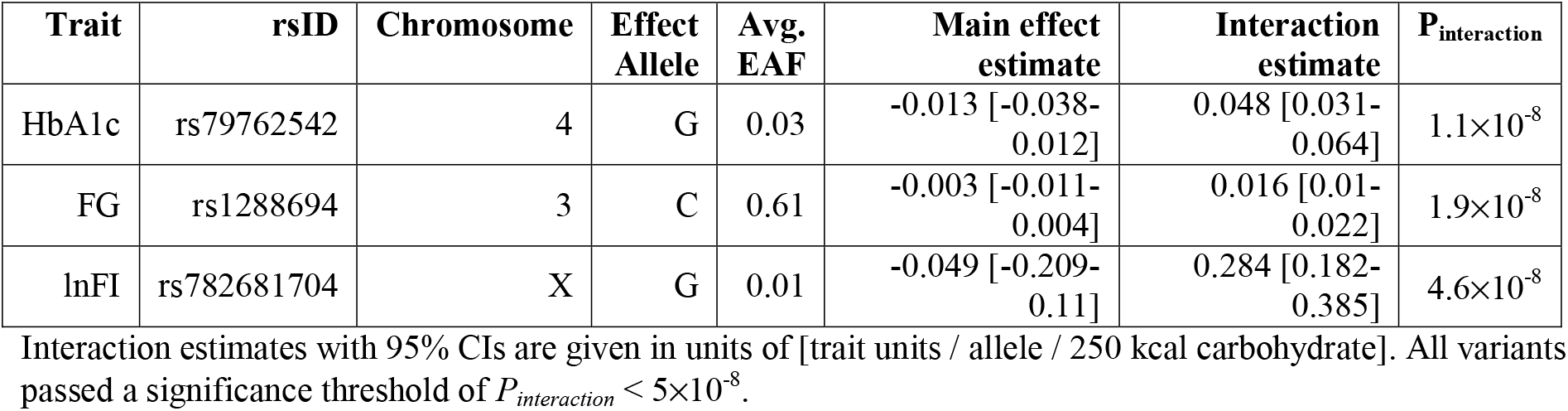
Top variants interacting with carbohydrate intake from the common-variant GWIS.

As the only variant reaching study-wide significance in the primary analysis, we looked deeper into the biological function of rs79762542 and the functional form of its interaction. Variant rs79762542 is observed on African-ancestry haplotypes and was discovered with respect to HbA1c. The variant does not have known regulatory activity based on epigenomic assays in RegulomeDB, but there is evidence for a role in regulating expression of the nearby gene *FRAS1*, especially in thyroid, where this gene is most strongly expressed (GTEx project). Colocalization analysis did not support a shared causal signal between our interaction results and thyroid-specific eQTL signal (posterior probability of shared causal variant = 0.003%).

In genotype-stratified meta-analysis, HbA1c showed a modest negative association with increasing carbohydrate relative to fat intake in major allele homozygotes (−0.033%HbA1c/250kcals; *p* = 0.004) versus a positive association in minor allele carriers (0.10%HbA1c/250kcals; *p* = 0.42) that may not have reached significance due to the much lower sample size in this group (N = 1,055 across all studies). This genetic effect modification was moderately consistent across the cohorts in which HbA1c levels were studied and the minor allele of rs79762542 was observed (Fig. 1a,b; identical visualization in the African American race/ethnicity subset in Supp. Fig. S4a,b). Finally, with respect to the other glycemic traits in our analysis, interaction effects were directionally consistent but did not reach nominal significance (0.02 mmol/L/allele/250kcals; *p* = 0.07 for FG and 0.003 log(pmol/L)/allele/250kcals; *p* = 0.74 for lnFI).

**Figure 1:**
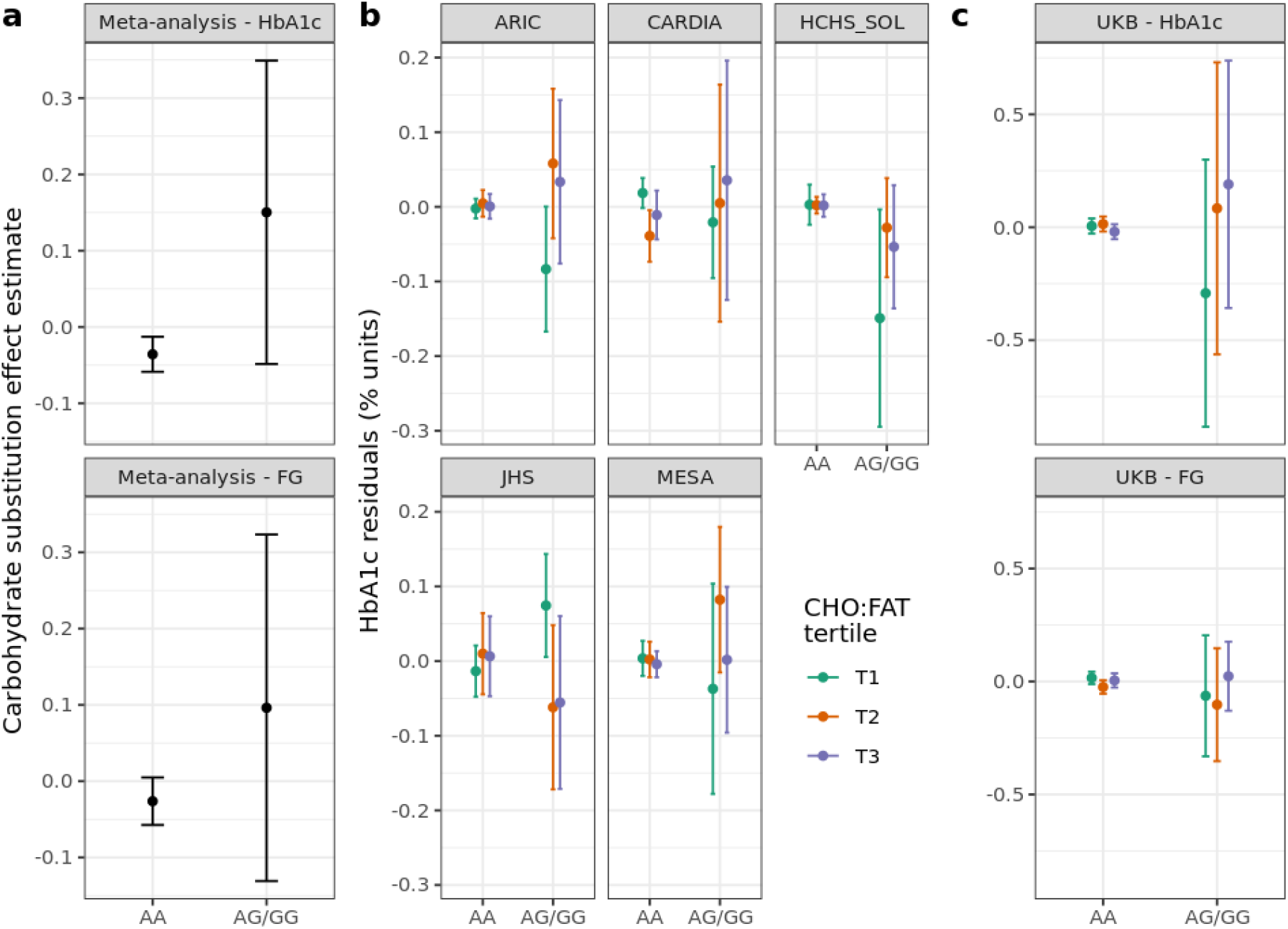
Exploration of the rs79762542 interaction and replication. (a) Genotype-stratified dietary main effect estimates. (b) Stratified plots (one for each cohort with HbA1c available) display residualized HbA1c within strata defined by both genotype at rs79762542 (none vs. any minor alleles) and cohort-specific tertile of carbohydrate:fat ratio. (c) Similar stratified plots for the UKB replication cohort. For (b) and (c), the *y*-axis displays residuals after regressing the relevant trait (HbA1c or FG) on the set of covariates used in the replication analysis.

Lookups for the other two variants passing *p* < 5×10^−8^ revealed potential functional roles for these variants. Variant rs1288694 (common in multiple ancestries) impacted FG in our analysis. The variant is intronic to the *FOXP1* gene and may regulate splicing of the same gene (GTEx project). *FOXP1* has a demonstrated role in hepatic gluconeogenesis(16). Variant rs782681704 is observed on African-ancestry haplotypes and was discovered with respect to FI in our analysis. The variant is intronic to *BRCC3* and has likely regulatory activity (RegulomeDB score of 0.59) but does not have clear evidence as an eQTL for *BRCC3*.

We explored these three prioritized single-variant loci through sensitivity analysis (Supp. Figs. S5 [rs79762542] and S6 [all three variants]). Interaction effects were robust in population subsets: only males, only females, and individuals without obesity. Exclusion of individuals either with or without prediabetes (beyond the predefined exclusion of individuals with diabetes) partially attenuated the interaction signal; this might be expected due to the removal of a substantial portion of the glycemic trait spectrum. Further, adjustment for either a diet quality score (AHEI-2010) or smoking status (along with their genotype interactions) did not meaningfully impact estimates. Interaction estimates were also generally consistent in the African American race/ethnicity subset, indicating that the interactions for African ancestry-specific variants do not solely reflect population stratification.

### Common variant replication

For the three prioritized single-variant loci, we tested for replication of these signals in the UK Biobank (N=178,352 with 24-hour dietary assessment data(17) and glycemic biomarkers; see Methods). Of these, 5,183 individuals were included in fasting glucose analyses (based on fasting for at least eight hours prior to the associated blood draw). In the full, multi-ancestry group (Supp. Table S5), we saw nominal replication of the interaction at rs79762542 with respect to both HbA1c (the discovery trait; *p* = 0.025) and fasting glucose (*p* = 0.013; Fig. 1c). Because most of the prioritized variants were specific to African-ancestry individuals, we conducted a similar replication in just this subgroup of the UK Biobank (Supp. Table S6). This analysis supported the rs79762542 interaction influencing FG (*p* = 0.046).

### Rare variant interactions

Rare variants (MAF < 1%) were analyzed in gene-centric, set-based tests, which help to overcome power limitations for low-MAF variants by aggregating signal across multiple variants annotated to the same gene. We used three variant aggregation strategies to define sets: selecting missense variants, loss-of-function variants, or a broader coding + non-coding variant set annotated to each gene (see Methods). No rare variant interaction signals showed genome-wide significance (*p* < 0.05 / 28,111 total genes = 1.78×10^−6^; Supp. Table S4; Supp. Fig. S7).

Since the set of rare variants used does not overlap with those from the common-variant tests, these gene-based tests can provide orthogonal evidence supporting common-variant signals while further clarifying potential effector genes. Each of the three prioritized single-variant findings were annotated to one or more genes based on proximity and/or expression-quantitative trait locus (eQTL) data. None of these pairings showed supporting gene-based signal for the corresponding glycemic trait, though the single study-wide significant variant (rs79762542, discovered in relation to HbA1c) showed a nominal corresponding signal from the gene-based test of *FRAS1* impacting FG (*p* = 0.028).

### Power calculations incorporating measurement error

Given the modest discovery of GDIs despite the use of the maximal sample available within TOPMed cohorts and substantial harmonization effort, we sought to better understand the necessary power to detect expected GDI effects. Simulation-based power calculations for single-variant tests were conducted with added noise to account for the known random measurement error in dietary data. Using literature-based anchors for expected effect sizes (Fig. 2a) and assuming a conservative but realistic dietary measurement reliability of 0.5 (see Methods for details), we established that a sample size of over 150,000 would be required to detect a GDI effect of 0.025 %HbA1c / allele / s.d. carbohydrate at genome-wide significance for a variant with MAF = 0.1 (Fig. 2b,c). As previously explored in the literature(18,19), power scaled approximately linearly with exposure measurement fidelity. If we alternatively assume perfect dietary exposure measurement, the associated sample size to detect the same effect was reduced to about 80,000 indicating the importance of accounting for this measurement error. The necessary sample size given realistic measurement reliabilities increased even further for lower-frequency variants (e.g., almost 1.2 million for MAF = 1%).

**Figure 2:**
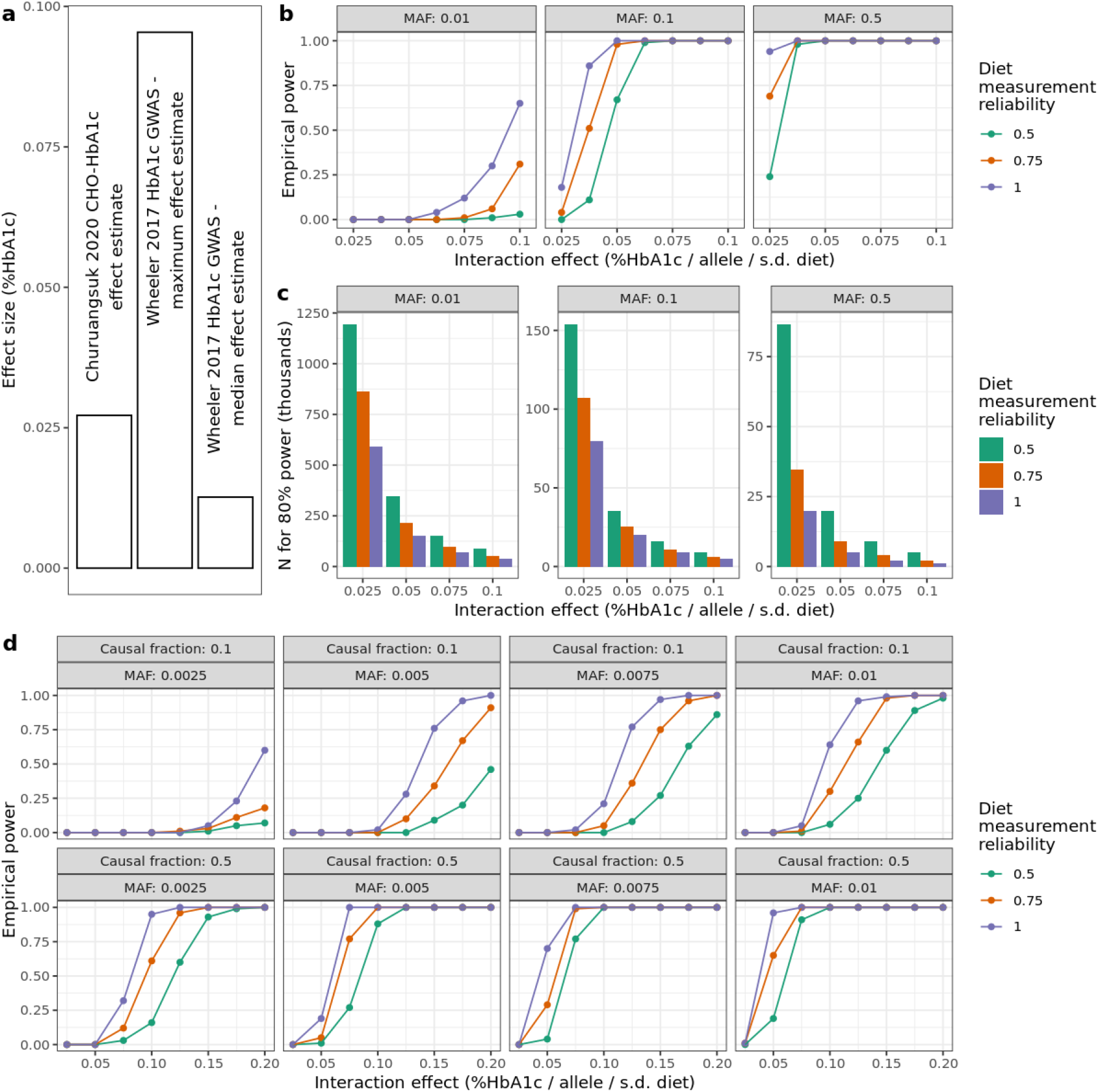
Power calculations for GEI incorporating exposure measurement error. For all plots above, HbA1c is used as a basis for parameter choices. (a) Genetic and dietary effect sizes on HbA1c for reference for potential interaction effects. (b) Simulation-based empirical power estimates are shown as a function of the interaction effect (*x*-axis), minor allele frequency (MAF; panels left-to-right), and diet measurement reliability (colors). (c) Bar plots show the estimated sample size needed to achieve 80% statistical power. Panels and colors are as in (b). (d) As in (a) but modeling empirical power for simulated aggregate tests of 20 rare variants with a causal fraction of 0.1 or 0.5 (indicated in panel labels). Additional assumptions for these simulations (full details in Methods): N = 35,000, phenotype mean of 5.5, phenotype standard deviation of 0.5, exposure mean of 0, exposure standard deviation of 1, genetic main effect of 0.01, and environmental main effect of 0.2.

We extended this simulation-based power calculation approach to test multiple variants jointly, mimicking the variant set-based test implemented for rare variants. Assuming similar measurement fidelity and effect sizes as for single variants and fixing the sample size to match the full sample used here (∼35,000), an aggregate test of 20 rare variants with a causal fraction of 50% and MAF = 0.25% had negligible power (Fig. 2d). Power increased somewhat but remained low when incorporating larger effect sizes (as are known to be present for rare variant main effects on cardiometabolic traits)(20). For example, using an effect size of 0.1, approximately equal to the largest genetic main effect on HbA1c reported by Wheeler and colleagues(21), power increased to 0.16.

## DISCUSSION

Our goal was to investigate genotype-related variability in the association of dietary macronutrient composition with glycemic traits. Importantly, this was based on a regression strategy modeling an isocaloric increase in dietary carbohydrate at the expense of fat(22). We conducted our comprehensive analyses in cohorts with racial/ethnic diversity with data for both common and rare variants from WGS. We examined multiple single variants with potential modifying roles for the relationship of carbohydrate intake with glycemic traits, but did not find substantial evidence from gene-based tests for a role of rare variants in modifying this diet-glycemia relationship. Furthermore, our simulation-based power analysis highlighted the impact of dietary measurement error on statistical power for the gene-diet interaction tests.

Dietary main effect models indicated that an increase in carbohydrate at the expense of dietary fat was associated with lower fasting glucose and HbA1c. The impact of this macronutrient exchange on glucose homeostasis and diabetes risk is complex and likely depends on the respective macronutrient quality. Prior studies suggest null associations of total carbohydrate to total fat exchange on diabetes risk(23,24). However, an exchange of animal-sourced fat for carbohydrate or vegetable fat appears to have favorable associations with HbA1c(25,26).

Our primary genome-wide common variant interaction analysis yielded an interaction between a 250kcal carbohydrate-fat substitution and HbA1c with the African-ancestry rs79762542 variant, which was validated in the UK Biobank. Genotype stratified analyses suggested that minor allele carriers generally had a small negative association between carbohydrate and HbA1c (−0.033%HbA1c/250kcals; *p* = 0.004) versus a larger but non-significant association in minor allele carriers where the sample size was much lower (−0.10 %HbA1c/250kcals; *p* = 0.42). However, this precise pattern was not observed in all cohorts. These results warrant further exploration in additional cohorts with African ancestry individuals and dietary intervention studies to examine whether glycemic traits among minor allele carriers may benefit from higher fat and lower carbohydrate diet composition. This primary discovery was made with HbA1c as an outcome, but our results from set-based rare variant analysis and the UK Biobank replication suggest similar patterns with respect to other traits such as FG. Beyond GDI discovery, the GWIS results provided an opportunity to evaluate the effect size assumptions used in the power calculations. For example, the rs79762542 interaction had an effect size of 0.048 %HbA1c/allele/250 kcal carbohydrate, or 0.068 %HbA1c/allele/std. dev. carbohydrate. This effect size is somewhat larger than, but of a similar magnitude to, the relevant anchor for the power analysis (the referenced main effect association of carbohydrate with HbA1c(27)).

This analysis leveraged WGS data along with multi-variant set-based tests to better incorporate rare variants (minor allele frequency [MAF] < 0.01). While these variants don’t contribute substantially to the overall population variance of glycemic or other traits, they tend to have larger effect sizes and thus may be important for the specific individuals carrying them(14). For example, phenylketonuria, a well-known inborn error of metabolism, acts through a rare-variant GDI in which severe adverse effects of phenylalanine intake are seen only in individuals with a particular genotype(28). In our study, despite helping to reinforce common variant signals, the rare variant analysis did not contribute additional findings after aggregation at the gene level. Substantially larger sample sizes will likely be necessary to uncover macronutrient GDIs involving rare variants.

We explored the statistical power for these interaction tests through simulations incorporating random dietary measurement error using available simulation-based power calculation software (ESPRESSO.GxE(29), for single variants) with additional extensions to allow for aggregate rare variant tests. We estimated that substantially higher sample sizes (almost five times that used in this study) are required for sufficient power to detect macronutrient-gene interactions at expected effect sizes obtained from genetics and nutrition literature. This prompts two directions of further inquiry. First, it suggests the importance of complementary approaches that assess where there is any whole-genome contribution to the diet-glycemia association, at least in observational datasets. These whole-genome analyses trade resolution for statistical power(30) and have a precedent for GDIs in smaller study samples(31). Second, it reinforces the importance of collecting dietary intake data in the growing group of large-scale biobanks and cohorts. Improvements in study design, data collection methods, and analysis that can improve quality of dietary assessments are also warranted. For example, conducting rigorous validation studies of the data collection tools and approaches and ascertaining repeated dietary data can greatly improve the precision of these measurements on a population level. Advancements in objectively quantifying habitual diet from biospecimen samples are also underway and have potential to improve discovery for genetic analyses.

An important strength of this study is the breadth of ethnic and cultural diversity of the sample (increasing the likelihood that findings are robust) and of genetic variation (with WGS data enabling exploration of ancestry-specific genetic variation across the frequency spectrum). We also conducted a systematic investigation into the available statistical power while incorporating both realistic degrees of measurement error and evidence-based estimations of realistic effect sizes for gene-macronutrient interactions. However, the diversity of the included study sample also introduces heterogeneity that may be problematic. For example, the cohorts used different dietary assessment tools to capture habitual intake, leading to differences in the degree and direction of random and systematic measurement error. This is compounded by general, culturally-driven differences in food intake across race/ethnicity groups. Further heterogeneity arises from the time of data collection; the perceptions of carbohydrate intake have trended as more and less healthful in recent decades, potential resulting in differential confounding between diet and other health-related behaviors depending on the time of data collection(32). Future work can step beyond broad macronutrient categories by harmonizing intakes of specific foods or dietary patterns and analyzing macronutrient subtypes (e.g., added sugar and specific fatty acids). These approaches, combined with improved methods for detecting rare variant gene-environment interactions, will help utilize the increasing volume of WGS data to discover new GDIs relevant for metabolic disease risk.

## METHODS

### Whole-genome sequencing

All genetic and phenotypic data used for this study were obtained from the NCBI Database of Genotypes and Phenotypes (dbGaP) and the research was approved under the Mass General Brigham IRB (protocol 2017P000531). WGS was conducted through the NHLBI TOPMed program (Freeze 8 data release). Sequencing and alignment to the GRCh38 reference genome was performed across six centers across the US: Broad Institute of MIT and Harvard, Northwest Genomics Center, New York Genome Center, Illumina Genomic Services, PSOMAGEN (formerly Macrogen), Baylor College of Medicine Human Genome Sequencing Center, McDonnell Genome Institute (MGI) at Washington University. Data harmonization and joint variant discovery and genotype calling were performed within the TOPMed Informatics Research Center at the University of Michigan. Sequence quality control filters were as follows: estimated DNA sample contamination below 10% and at least 95% of the genome having coverage of at least 10x. After genotyping, variants were further filtered for Mendelian inconsistency (based on a support vector machine classifier) and excess heterozygosity. Additional sample quality control was performed within the Data Coordinating Center at the University of Washington, including: matching sex as annotated and inferred from WGS, concordance of WGS genotypes with prior array-based “fingerprints”, and agreement of inferred relatedness with expectations based on pedigrees. Additional details on the processing steps are available at: https://www.nhlbiwgs.org/topmed-whole-genome-sequencing-methods-freeze-8.

Global measures of ancestry and relatedness were calculated on the entire TOPMed Freeze 8 sample by the TOPMed Data Coordinating Center. Genetic principal components reflecting ancestry were calculated using the PC-AiR method (allowing for related individuals)(33), and kinship matrices were calculated using the PC-Relate method (accounting for principal components)(34), both from the GENESIS R package. A sparse genetic relationship matrix containing only relationships of degree four or closer was extracted for analysis. Samples were grouped into race/ethnicity categories based on cohort-reported values.

### Harmonization of glycemic traits

Phenotypes were harmonized across the 10 studies based on a protocol developed within the TOPMed Diabetes Working Group. Glycemic traits, including fasting glucose (FG; mmol/L), fasting insulin (FI; pmol/L), and glycated hemoglobin (HbA1c; %), were collected where available. Fasting (for FG and FI) was defined as at least 8 hours without food or drink. FG measurements made in blood rather than plasma were adjusted by multiplying by a correction factor of 1.13. When multiple values were available for a given participant, blood draws were chosen to favor measurements made at study baselines and in order to maximize overlap with time points in which dietary data were collected. Participants were excluded if their glycemic trait blood draw was more than one year before or after diet measurement or if they had diabetes (defined as any of: taking anti-diabetic medication, FG ≥ 7 mmol/L, or HbA1c ≥ 6.5%). Further study-specific details are available in the Supplementary Methods. Phenotype data harmonization and all other post-genome-wide analyses and visualizations were conducted using R version 4.1.1(35). Unless otherwise noted, all analyses including harmonization were performed on the NHLBI BioData Catalyst cloud computing platform(36).

### Harmonization of dietary data

Estimates of dietary intake were derived from self-reported diet questionnaires, either food frequency questionnaires (FFQ), diet history, or 24-hour recalls (24HR). Reported quantities of food and beverage consumption were converted into daily nutrient intake estimates via standard nutrient databases (see Supplementary Methods for study-specific details), with energy and macronutrients (carbohydrate, protein and total fat) expressed in kilocalories/day (kcals/d). Participants were excluded if responses were deemed implausible, based on having total caloric intake <600 kcals/d or >4800 kcals/d. Nutrient intake values were analyzed in units of kcals/d and winsorized at three standard deviations from the mean. Dietary fiber was represented in grams/day (g/d) and alcohol intake was reported as number of drinks per day.

### Genome-wide gene-diet interaction scans

For each cohort and glycemic trait, four genome-wide gene-diet interaction scans were performed to identify diet-interacting loci: one for common variants, and three gene-based aggregate tests for rare variants using different variant masks (described below). Mixed linear models were used to allow for random effects of kinship capturing close family relationships (degree four relatives or closer). The linear model setup was as follows:

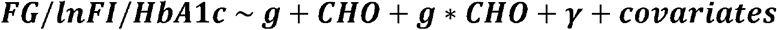

Where ***g*** is the genotype at the variant of interest, ***CHO*** is dietary carbohydrate intake (kcals/d), and ***γ*** is a random effect governed by a sparse kinship matrix. General covariates included sex, age, age^2^, five genetic principal components to capture genetic ancestry, cohort-reported race/ethnicity to capture potential confounding by ethnicity-related dietary behavior, and additional study-specific covariates (Supp. Table S2). Dietary protein intake and total energy (also expressed in kcals/d) were included as covariates to set up an isocaloric substitution model in which increases in CHO were implicitly exchanged for decreases in dietary fat. Dietary fiber (g/d), alcohol intake (standard drinks/d), and body mass index (BMI; kg/m^2^) were included as additional covariates to account for further lifestyle-related confounding. During null model fitting, heterogeneous variances were allowed within each cohort-reported race/ethnicity group (equivalent to including a random effect for this grouping variable).

Genome-wide interaction analysis was performed using the MAGEE package(37). Single-variant analysis (*glmm*.*gei* function) was conducted for variants with MAF > 1%. METAL(38) was used to perform fixed effects meta-analysis across cohorts. Specifically, the 2-degree of freedom joint meta-analysis patch was used(39), with genetic main effect and interaction p-values derived downstream based on the resulting effect and standard error estimates.

Gene-centric, set-based rare-variant analysis (*MAGEE* function) was conducted for variants with MAF < 1%. Variant annotations derived from the WGSA v0.8 and WGSAParsr v6.3.8 were retrieved from the NCBI dbGaP database. A genome-wide interaction meta-analysis was conducted for each of three variant masks: loss of function variants (VEP_ensembl_Consequence has terms *transcript_ablation, splice_acceptor_variant, splice_donor_variant, stop_gained, frameshift_variant, stop_lost, start_lost* or *transcript_amplification*), missense variants (VEP_ensembl_Consequence has the term *missense_variant*), and a broad coding and noncoding filter (containing high-confidence loss-of-function variants, missense variants, protein-altering variants, synonymous variants, variants overlapping enhancers, and variants overlapping promoters). *MAGEE* calculates three interaction p-values: an adjusted variance component-like test, a burden test (assuming a consistent direction of effect for all variants), and a hybrid test (which combines the first two p-values using Fisher’s method). P-values from the hybrid test were used here to balance the increased power of the burden test with the possibility that its assumption of homogeneous effect directions is violated. Meta-analysis was then performed using a fixed-effects strategy.

Linear mixed models without genotype terms, meant to understand the marginal dietary effects prior to considering genetic effects, were fit in R using analogous models to those with diet-genotype interaction terms. Diet-genetic principal component interaction terms were excluded from these models and individuals in cohort-reported race/ethnicity groups with less than five members were excluded. Fixed effect meta-analysis of the CHO association (implicitly modeling an exchange with FAT due to the additional dietary covariates) was conducted using the *meta* package.

### Variant follow-up

Sensitivity analysis was conducted to understand the impact of modeling choices on the interaction effect estimates derived in the genome-wide analysis. These linear mixed models were fit in R, with G×CHO interaction terms subject to fixed-effects meta-analysis using the *meta* package as with the models without genotype effects. Some of these involved subsets of the population: male and female subsets were tested separately, as well as subsets without obesity (BMI < 30 kg/m^2^) and with and without prediabetes (defined as FG > 5.6 mmol/L or HbA1c > 5.7%). Additional models included adjustment for either smoking status (never/former/current, coded as 0/1/2 and analyzed as a continuous variable), the Alternative Healthy Eating Index 2010(40) (a diet quality score), or a categorical coding of alcohol intake (none, modest [<1 drink/day for females or <2 drinks/day for males], or high), where available. These models with additional covariate adjustments also included adjustment for their interactions with genotype. Finally, a model including genotype interaction terms for other main dietary components and lifestyle confounders (total energy, protein, fiber, and alcohol) was included. This type of residual confounding by genotype-covariate interaction terms has been previously documented(41), but would have decreased statistical power if included in the genome-wide analysis, especially for lower-frequency variants.

Variant rs79762542 was investigated in greater depth as the only variant reaching study-wide significance. Based on its eQTL relationship impacting *FRAS1* gene expression in thyroid from the GTEx v8 dataset (https://gtexportal.org/), we tested for colocalization of this signal with the carbohydrate interaction signal impacting HbA1c. Interaction summary statistics were retrieved in a window of 1Mb around the index variant rs79762542, and all thyroid-specific *cis-*eQTL summary statistics related to *FRAS1* were retrieved from GTEx. Colocalization was tested using the *coloc* package for R, assuming a single causal variant (*coloc*.*abf* function).

### Replication analysis in the UK Biobank

UKB is a large prospective cohort with both deep phenotyping and molecular data, including genome-wide genotyping, on over 500,000 individuals ages 40-69 living throughout the UK between 2006-2010(42). Genotyping, imputation, and initial quality control on the genetic dataset have been described previously(43). Analyses were conducted on genetic data release version 3, with imputation to a joint reference panel including the Haplotype Reference Consortium and the 1000 Genomes Project (1KGP), under UK Biobank application 27892. This work was conducted under a Not Human Subjects Research determination (NHSR-4298 at the Broad Institute of MIT and Harvard).

Ancestry group labels, genetic principal components, and labels defining an unrelated subset of individuals were retrieved from the Pan-UKBB project (https://pan.ukbb.broadinstitute.org/; data retrieved from UKB return of results number 2442). Only unrelated individuals were used for analysis, with additional removal of individuals who were pregnant or had diabetes at the study center visit. Two glycemic traits were available for testing in UKB: HbA1c and glucose (collected as a random glucose measurement and later to remove non-fasting individuals). Outliers for both traits (defined as more than 5 standard deviations from the mean) were removed. Dietary data came from one or more Oxford WebQ 24-hour dietary assessments(17) completed at the study center or during online follow-up over the course of approximately two years. Daily nutrient intake estimates (calculated centrally by the UKB) were averaged across all questionnaires for each individual and winsorized at three standard deviations from the mean. After all exclusions, 178,352 individuals without diabetes had available genotype, biomarker, and dietary data.

Regression analysis in the UKB mirrored that of the primary analysis, replacing cohort-reported race/ethnicity with genetically-defined ancestry groups as defined by the Pan-UKBB project. Given the larger sample size available, gene-covariate interactions were included for dietary covariates (total energy, protein, fiber, and alcohol). When analyzing glucose, only the subset of individuals with reported fasting times of at least eight hours were included, reducing the sample size to 5,183. Due to the African ancestry-specificity of some of the top variants, a second replication analysis was performed in the African-ancestry subset of UKB.

### Power Calculations

Interaction test power calculations were performed using the *ESPRESSO*.*GxE* R package, which uses a simulation-based approach to calculate empirical power estimates (given some sample size) and sample size requirements (to achieve 80% power). The following parameters were fixed for this analysis, chosen to mimic an analysis of HbA1c: random seed = 1, significance threshold = 5×10^−8^, phenotype mean = 5.5, phenotype standard deviation = 0.5, phenotype reliability = 1, genetic main effect = 0.1, exposure mean = 0, exposure standard deviation = 1, exposure main effect = 0.2. The following parameters were varied: interaction effect {0.025, 0.0375, 0.05, 0.0625, 0.075, 0.0875, 0.1}, MAF {0.01, 0.05, 0.1, 0.5}, and exposure reliability {0.25, 0.5, 0.75, 1}. Here, reliability is used to quantify the simulated measurement error of the phenotype and exposure and is equivalent to an intraclass correlation coefficient (ratio of between-subject variance to total [between-subject plus measurement error] variance).

To enable simulation-based power calculations for aggregate tests of rare variants while accounting for exposure measurement error, we developed an extension of the *ESPRESSO*.*GxE* package, called *ESPRESSO*.*GxE*.*RV*. In this extension, the basic structure of the simulations remains the same, but an additional parameter allows the user to specify a number of variants (M) to test in aggregate. Within each simulation run, M variants are simulated, with some portion having equal interaction and main effects on the outcome (according to a user-specified causal variant fraction) and the rest generated randomly. The final p-value from that simulation is calculated using Fisher’s method on the full set of M p-values. The following parameters were given different values for this set of simulations: interaction effect {0.025, 0.05, 0.075, 0.1, 0.125, 0.15, 0.175, 0.2}, MAF {0.0025, 0.005, 0.0075, 0.01}. Other parameters were specific to rare variant tests: number of variants per aggregate test {1, 5, 10, 20} and causal fraction {0.1, 0.25, 0.5, 1}. Code for this extension of the package can be found on GitHub: https://github.com/kwesterman/ESPRESSO.GxE.RV.

To provide context for realistic gene-diet interaction effect sizes despite few well-replicated examples of such interactions for glycemic traits in the literature, we retrieved results from variants reaching significance in a recent trans-ancestry GWAS for HbA1c(21) and an estimated effect for the carbohydrate-HbA1c relationship from a recent nutritional epidemiological analysis(27).

## Data Availability

Data analyzed in the manuscript can be accessed via the NCBI Database of Genotypes and Phenotypes (dbGaP) pending application and approval from the NHLBI Trans-Omics for Precision Medicine (TOPMed) program.

## AUTHOR CONTRIBUTIONS

KEW, MEW, JM, and AKM designed the research. KEW, SMG, JW, DKT, and AKM contributed to the harmonization of glycemic trait variables across studies; KEW conducted the research and performed the primary data analysis; JCF, JBM, DKT, HC, and AKM supervised the research; KEW and MEW wrote the manuscript; all additional authors contributed to the collection and curation of the study-specific or TOPMed-wide datasets; KEW and AKM had primary responsibility for the final content. All authors read and approved the final manuscript: KEW, MEW, SMG, JW, DD, JM, AA, SA, ASB, AGB, MLB, JAB, YIC, JD, MOG, XG, NRH, AH, BH, MRI, WCJ, RRK, LL, RNL, CL, SL, JM, RN, JSP, MP, LR, LJR, ES, MKS, AHS, AVS, NLS, LS, ST, KDT, PSD, JGW, ACW, LRY, JY, YZ, EB, ACM, MF, TPR, BMP, DL, NLH, VSR, RAM, DKA, RK, KEN, AC, AC, JR, SSR, JEM, APR, CK, JCF, JBM, JM, DKT, HC, AKM.

## ACKNOWLEDGMENTS

The Genotype-Tissue Expression (GTEx) Project was supported by the Common Fund of the Office of the Director of the National Institutes of Health, and by NCI, NHGRI, NHLBI, NIDA, NIMH, and NINDS. The data used for the analyses described in this manuscript were obtained fromthe GTEx Portal on 03/28/2022.

Support for this work was provided by the National Institutes of Health, National Heart, Lung, and Blood Institute, through the BioData Catalyst program (award 1OT3HL142479-01, 1OT3HL142478-01, 1OT3HL142481-01, 1OT3HL142480-01, 1OT3HL147154-01). Any opinions expressed in this document are those of the author(s) and do not necessarily reflect the views of NHLBI, individual BioData Catalyst team members, or affiliated organizations and institutions. Further, the authors wish to acknowledge the contributions of the consortium working on the development of the NHLBI BioData Catalyst ecosystem.

The authors would also like to acknowledge the contributions of L. Adrienne Cupples to the Framingham Heart Study and the TOPMed program.

## TOPMed Acknowledgments

Molecular data for the Trans-Omics in Precision Medicine (TOPMed) program was supported by the National Heart, Lung and Blood Institute (NHLBI). See the TOPMed Omics Support Table below for study specific omics support information. Core support including centralized genomic read mapping and genotype calling, along with variant quality metrics and filtering were provided by the TOPMed Informatics Research Center (3R01HL-117626-02S1; contract HHSN268201800002I). Core support including phenotype harmonization, data management, sample-identity QC, and general program coordination were provided by the TOPMed Data Coordinating Center (R01HL-120393; U01HL-120393; contract HHSN268201800001I). We gratefully acknowledge the studies and participants who provided biological samples and data for TOPMed.

| Study | Acknowledgements |
| --- | --- |
| Atherosclerosis Risk in Communities study | ARIC: The Atherosclerosis Risk in Communities study has been funded in whole or in part with Federal funds from the National Heart, Lung, and Blood Institute, National Institutes of Health, Department of Health and Human Services, under Contract nos. (75N92022D00001, 75N92022D00002, 75N92022D00003, 75N92022D00004, 75N92022D00005). The authors thank the staff and participants of the ARIC study for their important contributions.<br>WGS for “NHLBI TOPMed: Atherosclerosis Risk in Communities (ARIC)” (phs001211) was performed at the Baylor College of Medicine Human Genome Sequencing Center (HHSN268201500015C and 3U54HG003273-12S2) and the Broad Institute for MIT and Harvard (3R01HL092577- 06S1).<br>The Genome Sequencing Program (GSP) was funded by the National Human Genome Research Institute (NHGRI), the National Heart, Lung, and Blood Institute (NHLBI), and the National Eye Institute (NEI). The GSP Coordinating Center (U24 HG008956) contributed to cross program scientific initiatives and provided logistical and general study coordination. The Centers for Common Disease Genomics (CCDG) program was supported by NHGRI and NHLBI, and whole genome sequencing was performed at the Baylor College of Medicine Human Genome Sequencing Center (UM1 HG008898). |
| CARDIA | The Coronary Artery Risk Development in Young Adults Study (CARDIA) is conducted and supported by the National Heart, Lung, and Blood Institute (NHLBI) in collaboration with the University of Alabama at Birmingham (HHSN268201800005I & HHSN268201800007I), Northwestern University (HHSN268201800003I), University of Minnesota (HHSN268201800006I), and Kaiser Foundation Research Institute (HHSN268201800004I). CARDIA was also partially supported by the Intramural Research Program of the National Institute on Aging (NIA) and an intra-agency agreement between NIA and NHLBI (AG0005). |
| CHS | This research was supported by contracts 75N92021D00006, HHSN268201200036C, HHSN268200800007C, HHSN268201800001C, N01HC55222, N01HC85079, N01HC85080, N01HC85081, N01HC85082, N01HC85083, N01HC85086, and grants R01HL120393, U01HL080295 and U01HL130114 from the National Heart, Lung, and Blood Institute (NHLBI), with additional contribution from the National Institute of Neurological Disorders and Stroke (NINDS). Additional support was provided by R01AG023629 from the National Institute on Aging (NIA). A full list of principal CHS investigators and institutions can be found at CHS-NHLBI.org. The content is solely the responsibility of the authors and does not necessarily represent the official views of the National Institutes of Health. |
| Framingham Heart Study | WGS for "NHLBI TOPMed: Whole Genome Sequencing and Related Phenotypes in the Framingham Heart Study (phs000974.v1.p1) was performed at the Broad Institute of MIT and Harvard (3R01HL092577-06S1 (AFGen)). This research was conducted in part using data and resources from the Framingham Heart Study of the National Heart Lung and Blood Institute of the National Institutes of Health and Boston University School of Medicine. The Framingham Heart Study (FHS) acknowledges the support of contracts NO1-HC-25195, HHSN268201500001I and 75N92019D00031 from the National Heart, Lung and Blood Institute and grant supplement R01 HL092577-06S1 for this research. We also acknowledge the dedication of the FHS study participants without whom this research would not be possible. |
| GeneSTAR | GeneSTAR was supported by grants from the National Institutes of Health/National Heart, Lung, and Blood Institute (U01 HL72518, HL087698, HL49762, HL59684, HL58625, HL071025, HL092165, HL099747, K23HL105897, HL112064), by grants from the National Institutes of Health/National Institute of Nursing Research (NR0224103, NR008153), by a grant from the National Institutes of Health/National Institute of Neurological Disorders and Stroke (NS062059), by a grant from the National Institutes of Health/National Center for Research Resources (M01-RR000052) to the Johns Hopkins General Clinical Research Center, and by a grant from the National Center for Advancing Translational Sciences, National Institutes of Health (UL1TR001079) to the Johns Hopkins Institute for Clinical & Translational Research. |
| GOLDN | GOLDN biospecimens, baseline phenotype data, and intervention phenotype data were collected with funding from National Heart, Lung and Blood Institute (NHLBI) grant U01 HL072524. Whole-genome sequencing in GOLDN was funded by NHLBI grant R01 HL104135-04S1. We thank GOLDN participants and investigators for their significant contributions. |
| HCHS/SOL | The Hispanic Community Health Study/Study of Latinos is a collaborative study supported by contracts from the National Heart, Lung, and Blood Institute (NHLBI) to the University of North Carolina (HHSN268201300001I / N01-HC-65233), University of Miami (HHSN268201300004I / N01-HC-65234), Albert Einstein College of Medicine (HHSN268201300002I / N01-HC-65235), University of Illinois at Chicago – HHSN268201300003I / N01-HC-65236 Northwestern Univ), and San Diego State University |
|  | (HHSN268201300005I / N01-HC-65237). The following Institutes/Centers/Offices have contributed to the HCHS/SOL through a transfer of funds to the NHLBI: National Institute on Minority Health and Health Disparities, National Institute on Deafness and Other Communication Disorders, National Institute of Dental and Craniofacial Research, National Institute of Diabetes and Digestive and Kidney Diseases, National Institute of Neurological Disorders and Stroke, NIH Institution-Office of Dietary Supplements. |
| JHS | The Jackson Heart Study is supported and conducted in collaboration with Jackson State University (HHSN268201800013I), Tougaloo College (HHSN268201800014I), the Mississippi State Department of Health (HHSN268201800015I) and the University of Mississippi Medical Center (HHSN268201800010I, HHSN268201800011I and HHSN268201800012I) contracts from the National Heart, Lung, and Blood Institute (NHLBI) and the National Institute on Minority Health and Health Disparities (NIMHD). The authors thank the participants and data collection staff of the Jackson Heart Study. The views expressed in this manuscript are those of the authors and do not necessarily represent the views of the National Heart, Lung, and Blood Institute; the National Institutes of Health; or the U.S. Department of Health and Human Services. |
| MESA | Whole genome sequencing (WGS) for the Trans-Omics in Precision Medicine (TOPMed) program was supported by the National Heart, Lung and Blood Institute (NHLBI). WGS for “NHLBI TOPMed: Multi-Ethnic Study of Atherosclerosis (MESA)” (phs001416.v1.p1) was performed at the Broad Institute of MIT and Harvard (3U54HG003067-13S1). Centralized read mapping and genotype calling, along with variant quality metrics and filtering were provided by the TOPMed Informatics Research Center (3R01HL-117626-02S1). Phenotype harmonization, data management, sample-identity QC, and general study coordination, were provided by the TOPMed Data Coordinating Center (3R01HL-120393-02S1), and TOPMed MESA Multi-Omics (HHSN268201500003I/HSN26800004). The MESA projects are conducted and supported by the National Heart, Lung, and Blood Institute (NHLBI) in collaboration with MESA investigators. Support for the Multi-Ethnic Study of Atherosclerosis (MESA) projects are conducted and supported by the National Heart, Lung, and Blood Institute (NHLBI) in collaboration with MESA investigators. Support for MESA is provided by contracts 75N92020D00001, HHSN268201500003I, N01-HC-95159, 75N92020D00005, N01-HC-95160, 75N92020D00002, N01-HC-95161, 75N92020D00003, N01-HC-95162, 75N92020D00006, N01-HC-95163, 75N92020D00004, N01-HC-95164, 75N92020D00007, N01-HC-95165, N01-HC-95166, N01-HC-95167, N01-HC-95168, N01-HC-95169, UL1-TR-000040, UL1-TR-001079, UL1-TR-001420, UL1TR001881, DK063491, and R01HL105756. The authors thank the other investigators, the staff, and the participants of the MESA study for their valuable contributions. A full list of participating MESA investigators and institutes can be found |
|  | at http://www.mesa-nhlbi.org. Also supported in part by the NHLBI contracts R01HL151855 and R01HL146860. |
| WHI | The WHI program is funded by the National Heart, Lung, and Blood Institute, National Institutes of Health, U.S. Department of Health and Human Services through contracts 75N92021D00001, 75N92021D00002, 75N92021D00003, 75N92021D00004, 75N92021D00005. |

Data described in the manuscript will be made available upon request pending application and approval from NHLBI Trans-Omics for Precision Medicine (TOPMed) program.

