## Supplementary Figures for "Investigating gene-diet interactions impacting the association between macronutrient intake and glycemic traits"

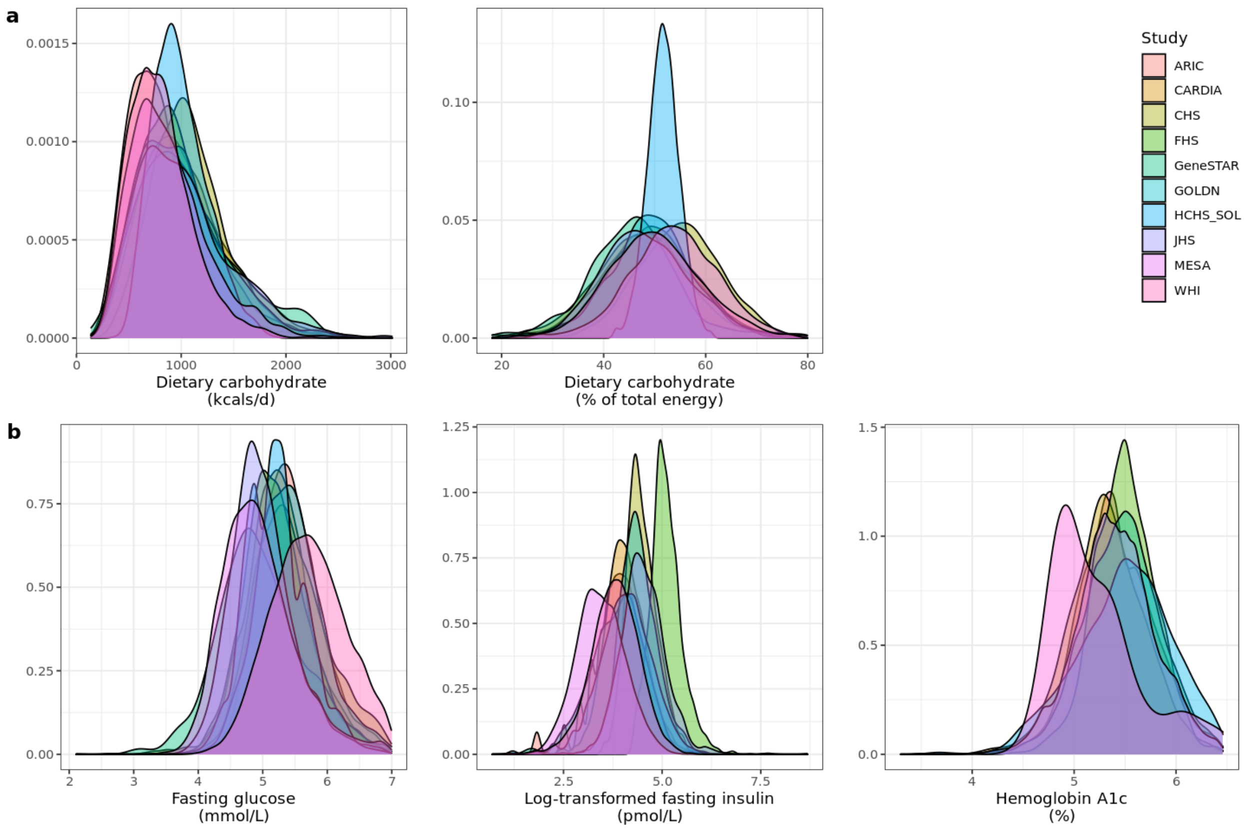


**Supplementary Figure S1:** Density plots illustrate the cohort-specific distributions of (a) carbohydrate intake, expressed as either kcals/day or percentage of total caloric intake, and (b) the three glycemic traits. We note that HCHS SOL was the only cohort whose dietary data were derived from 24-hour recalls rather than food frequency questionnaires.

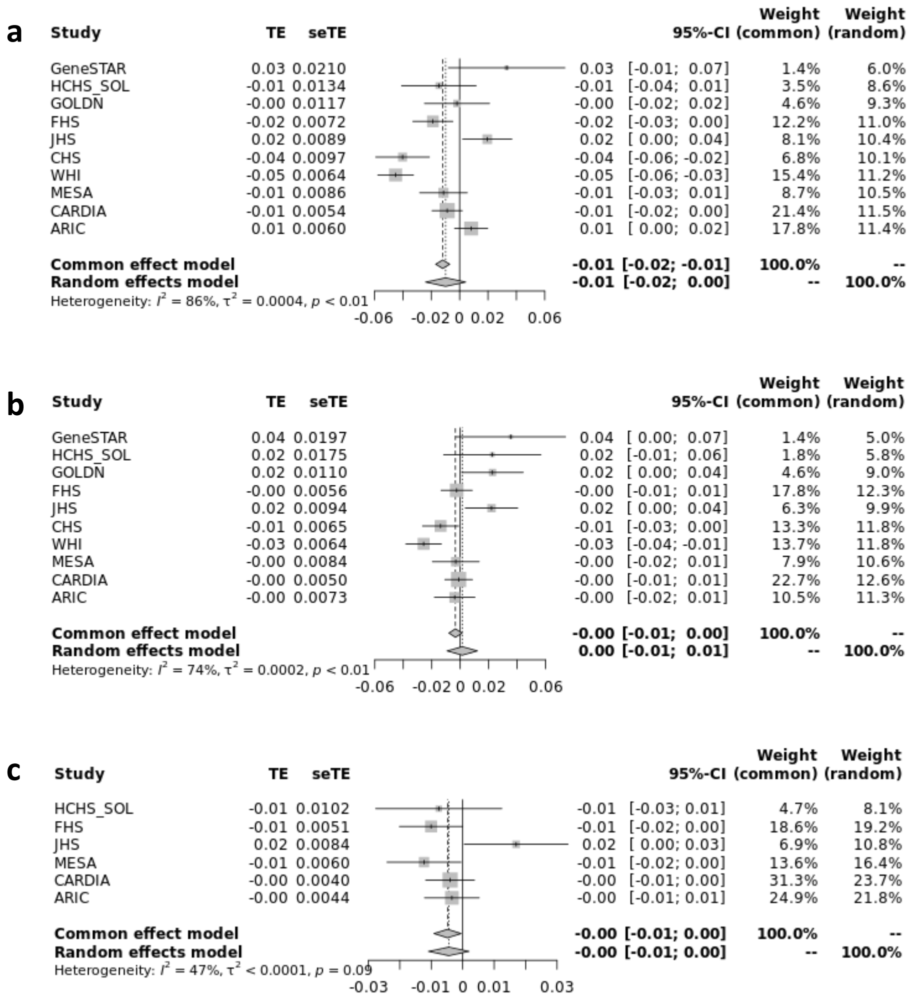


**Supplementary Figure S2:** Forest plots illustrating meta-analysis results for the association of a 100 kcal carbohydrate-fat exchange with each of: FG (a), lnFI (b), and HbA1c (c).

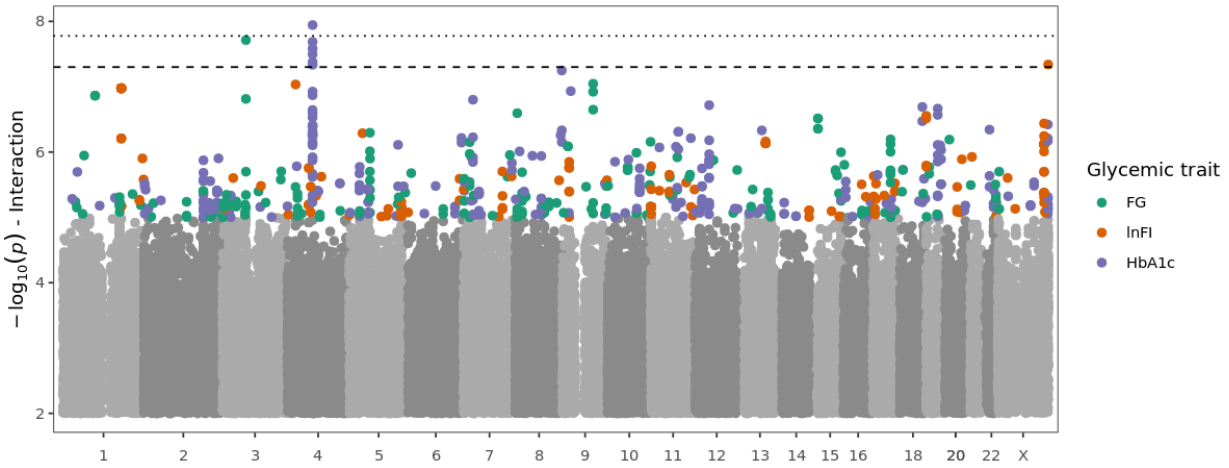


**Supplementary Figure S3:** Manhattan plot with single-variant interaction testing results for all glycemic traits. For each glycemic trait (colors, only shown for variants with $p < 10^{-5}$), -$\log_{10}$($p$-values) ($y$-axis) are shown as a function of genomic position ($x$-axis). The dashed line denotes a standard genome-wide significance threshold of $5{\times}10^{-8}$, and the dotted line denotes a study-wide significance threshold of $5{\times}10^{-8}$ / 3 = $1.67{\times}10^{-8}$. Only variants with $p < 0.01$ are shown.

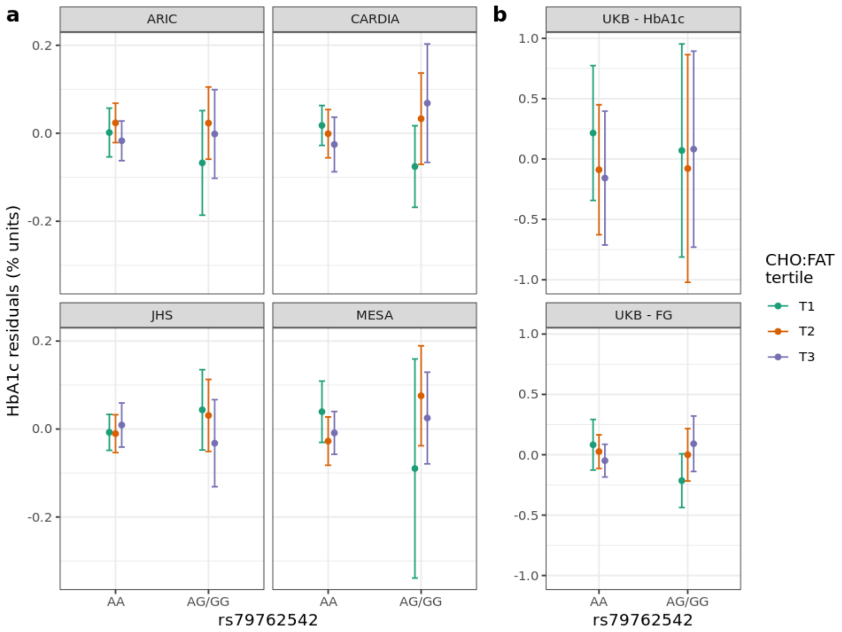


**Supplementary Figure S4:** Exploration of the rs79762542 interaction and replication in the population subsets with African race/ethnicity (TOPMed, [a]) and ancestry (UKB, [b]). (a) Stratified plots (one for each cohort with HbA1c available) display mean HbA1c within strata defined by both genotype at rs79762542 (none vs. any minor alleles) and cohort-specific tertile of carbohydrate:fat ratio. (b) Similar stratified plots for the UKB replication cohort. The $y$-axis displays residuals after regressing the relevant trait (HbA1c or FG) on the set of covariates used in the replication analysis.

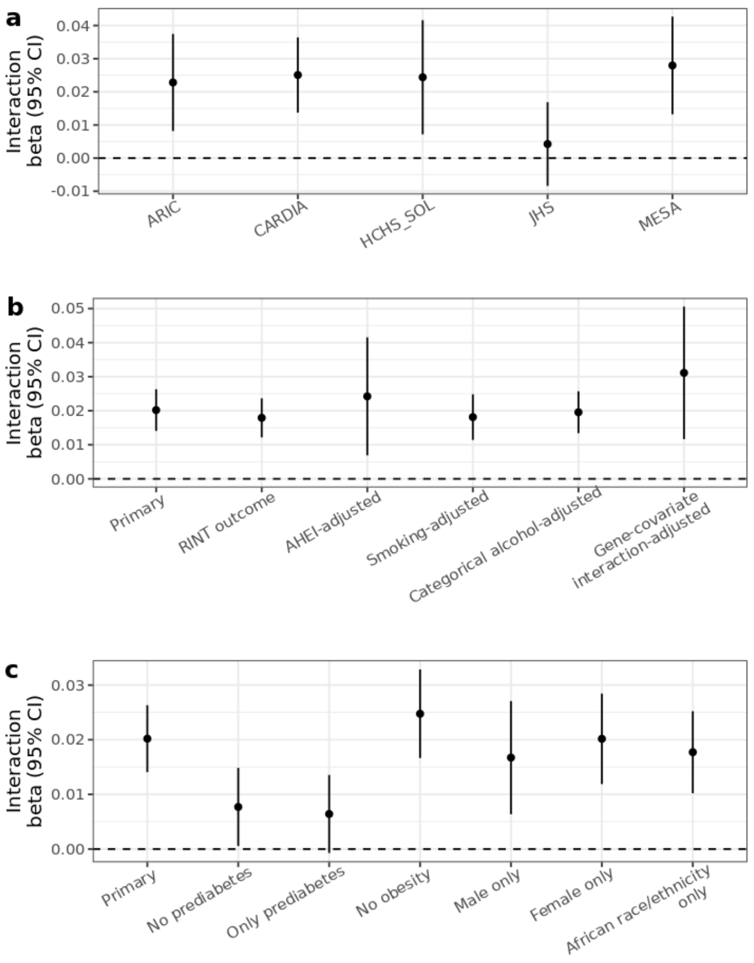


**Supplementary Figure S5:** Sensitivity analysis results for the study-wide significant variant rs79762542. (a) Cohort-specific interaction effect estimates for the raw HbA1c trait. (b) Interaction effect estimates from cross-cohort meta-analysis using alternative outcome transformations (RINT outcome) or additional covariate adjustments. Adjustments for AHEI, smoking, and categorical alcohol included their genotype-covariate interactions, and the "gene-covariate interaction-adjusted" model includes terms for total energy, kcals from protein, fiber intake, and alcohol intake. Categorical alcohol was coded as "none", "moderate" (less than one drink per day for women or two for men), or "high". (c) As in (b), but using varying subsets of the full cohorts rather than varying statistical adjustments. Prediabetes was defined as FG > 5.6 mmol/L or HbA1c > 5.7%. Obesity was defined as BMI > 30 kg/$m^2$.

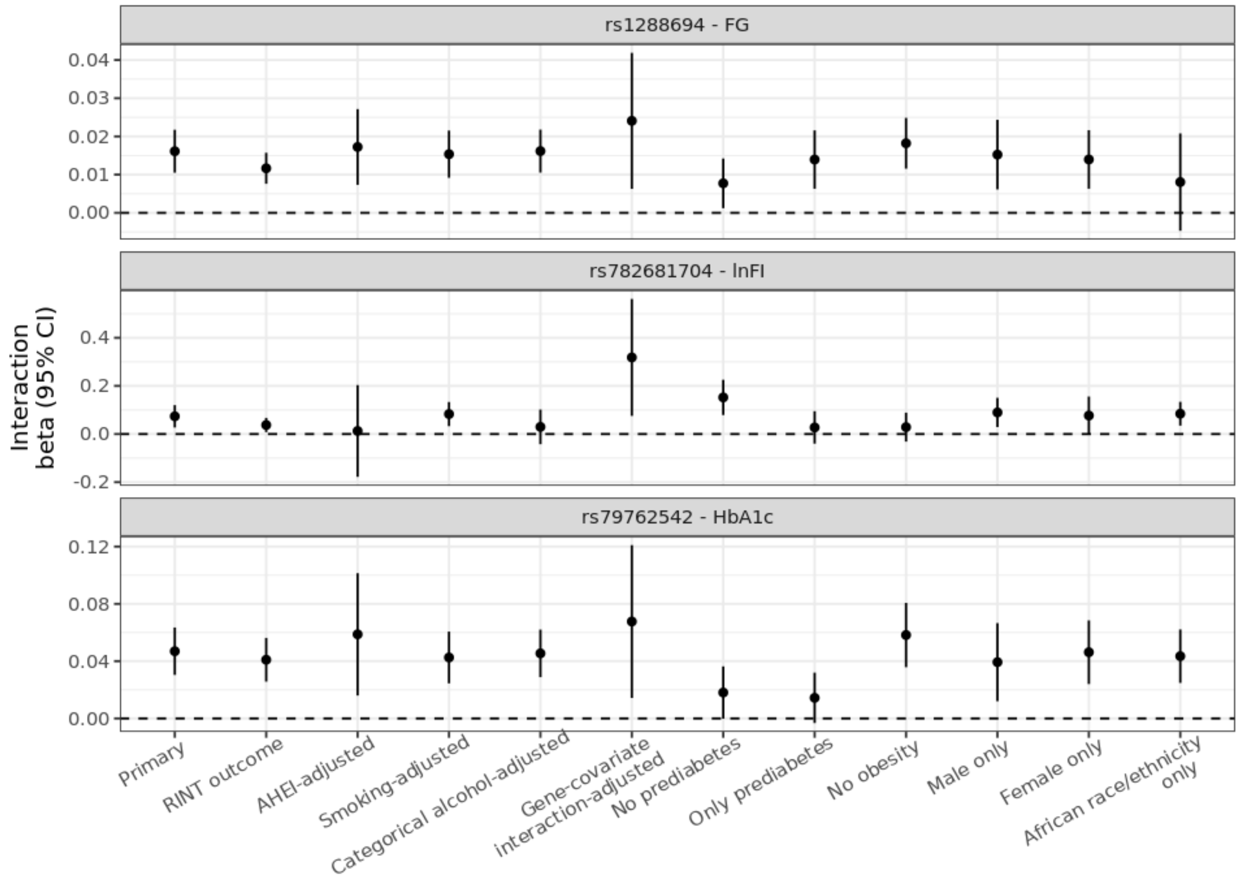


**Supplementary Figure S6:** Sensitivity analysis results for the three variants with $p < 5 \times 10^{-8}$. Panel headers indicate the tested variant and the associated glycemic trait. Adjustments and population subsets are defined as in Supp. Fig. S5.

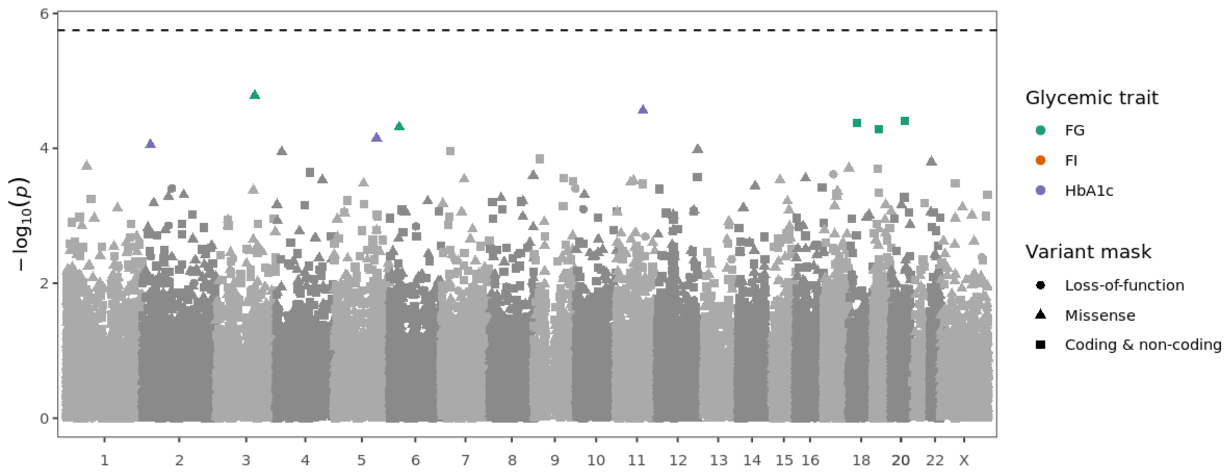


**Supplementary Figure S7:** Manhattan plot with aggregate rare-variant interaction testing results for all glycemic traits. For each glycemic trait (colors, only shown for variants with $p < 10^{-4}$), gene-based -$\log_{10}$($p$-values) ($y$-axis) are shown as a function of the average genomic position for variants included in each gene unit (*x*-axis). The dashed line denotes a Bonferroni significance threshold of 0.05 / 28,111 genes = $1.78\times10^{-6}$.
