## Supplementary Methods for "Investigating gene-diet interactions impacting the association between macronutrient intake and glycemic traits"

Notes on study-specific study design, protocols, exclusion criteria, or data processing are described below.

*ARIC*

The Atherosclerosis Risk in Communities (ARIC) study began in 1987 enrolling almost 16,000 Black and White adults aged 45-64 years who lived near one of the four field centers located in Jackson, MS, Minneapolis, MN, Washington County, MD, and Forsyth County, NC. Study participants were invited to take part in study visits for measurement of cardiovascular risk factors, medical history, dietary intake, physical activity, and other measures. Dietary intake (assessed by a modified Willett 66-item food frequency questionnaire) and FG and FI measurements were available at Visit 1 (1987-1989), while HbA1c measurements were available only at Visit 2 (1990-1992). Thus, there is a gap of approximately three years between the dietary and glycemic trait measurements for only the HbA1c analyses in ARIC.

*CARDIA*

The Coronary Artery Risk Development in Young Adults (CARDIA) study began in 1985 enrolling 5,115 Black and White men and women aged 18-30 years who lived in Birmingham, AL, Chicago, IL, Minneapolis, MN or Oakland, CA. The objective of this study was to better understand the etiology of the development of cardiovascular disease over many years of followup. Dietary intake, assessed by a diet history questionnaire, as well as FG and FI measures were available at baseline.

*CHS*

The Cardiovascular Health Study (CHS) is a population-based cohort study initiated by the National Heart, Lung and Blood Institute (NHLBI) in 1987 to determine the risk factors for development and progression of cardiovascular disease (CVD) in older adults, with an emphasis on subclinical measures. The study recruited 5,888 adults aged 65 or older at entry in four U.S. communities and conducted extensive annual clinical exams between 1989-1999 along with semi-annual phone calls, events adjudication, and subsequent data analyses and publications. Additional data are collected by studies ancillary to CHS. In June 1990, four Field Centers (Sacramento, CA; Hagerstown, MD; Winston-Salem, NC; Pittsburgh, PA) completed the recruitment of 5201 participants. Between November 1992 and June 1993, an additional 687 African Americans were recruited using similar methods. Blood samples were drawn from all participants at their baseline examination and during follow-up clinic visits and DNA was subsequently extracted from available samples. CHS analyses were limited to participants with available DNA who consented to genetic studies. The baseline examinations consisted of a home interview and a clinic examination that assessed not only traditional risk factors but also measures of subclinical disease, including carotid ultrasound, echocardiography, electrocardiography, and pulmonary function. Between enrollment and 1998-99, participants were seen in the clinic annually, and contacted by phone at 6-month intervals to collect information about hospitalizations and potential cardiovascular events. Major exam components were repeated during annual follow-up examinations through 1999. The study was initially approved by institutional review boards at the Field Centers (Wake Forest, University of California – Davis, Johns Hopkins University, University of Pittsburgh), the Core Laboratory (University of Vermont) and at the Coordinating Center (University of Washington). The University of Washington now handles CHS Data Repository approvals.

*GeneSTAR*

Genetic Studies of Atherosclerosis Risk (GeneSTAR) began in 1982 as the Johns Hopkins Sibling and Family Heart Study, a prospective longitudinal family-based study conducted originally in healthy adult siblings of people with documented early onset coronary disease under 60 years of age. Commencing in 2003, the siblings, their offspring, and the coparent of the offspring who were free of cardiovascular disease participated in a 2 week trial of aspirin 81 mg/day with pre and post ex vivo platelet function assessed using multiple agonists. Of the 2142 participants with pre- and post-aspirin measures of platelet function, 1683 were selected for TOPMed based on (1) participation in later GeneSTAR studies and (2) largest family size; 103 African American participants with the additional requirement of having coronary calcium measurements were later added, for a total of 1786 TOPMed GeneSTAR participants.

GeneSTAR participants were excluded based on any of: 1) the presence of any coronary heart disease or vascular thrombotic event, 2) pregnancy 3) serious medical disorders, including autoimmune diseases, renal or hepatic failure, cancer or HIV-AIDS, 4) chronic or acute use of glucocorticosteroid therapy, and 5) inability to independently make a decision to participate. T2D was defined as FG $\boldsymbol{\geq}$ 7 mmol/L, treatment for diabetes, physician diagnosis of diabetes, or self-reported diabetes. Glucose was measured in serum after a minimum 8 hour fast using a hexokinase assay on a Roche Diagnostics Modular DP chemistry analyzer. The inter-assay SD was 2.5 mg/dl (CV 2.9%) at 86 mg/dl. Insulin was measured from serum after a minimum 8 hour fast using an immunometric assay (DPC). The inter-assay SD was 1.2 uU/ml (CV 8.6%) at 14 uU/ml.

All nutrient and food servings were assessed with the Block Food Frequency Questionnaire at the baseline study visit.

*HCHS SOL*

The Hispanic Community Health Study / Study of Latinos (HCHS/SOL) is a community-based epidemiologic study of US Hispanic/Latino population from Bronx, Chicago, Miami, and San Diego. During 2008-2011, about 16000 participants (aged 18-74 years) were enrolled via a two-stage sampling (PMID: 20609343 and PMID: 20609344). First, census block group units were sampled at random with stratification based on Hispanic/Latino concentration and proportion of high/low socio-economic status in defined geographic areas. Then, in the second stage households were sampled at random, with stratification, from within selected census block groups. From household rosters, individuals in the 45-74 age group were oversampled. At baseline (2008-2011) and the second visit (2014-2017), the study participants underwent extensive clinic exams and assessments.

Dietary intake was assessed by two 24hr dietary recalls at baseline using the Nutrition Data System for Research software (Nutrition Data System for Research (NDS-R) version 2011. NDS-R software, 1998-1999). The first dietary recall was administered in-person at the field center and the second recall was administered via telephone at least 5 days and ideally within 45 days after the initial recall. The usual intake was predicted using the National Cancer Institute (NCI) method (PMID: 17000190). We used MIXTRAN and INDIVINT macros version 2.1 of the NCI method (https://epi.grants.cancer.gov/diet/usualintakes/macros_single.html) after excluding unreliable recalls according to the interviewer or extreme total energy intake (recall-sex specific below the 1^st^ percentile or above the 99th percentile). The model included covariates such as age, sex, Hispanic/Latino heritage, field center, weekday/weekend, and self-reported total intake amount (more, same, or less than usual amount).

Glucose (at least 8 hrs of fasting) was measured in EDTA plasma on a Roche Modular P

Chemistry Analyzer (Roche Diagnostics Corporation) using a hexokinase enzymatic method (Roche Diagnostics, Indianapolis, IN 46250). Insulin (at least 8 hrs of fasting) was measured in serum using a sandwich immunoassay method on a Roche Elecsys 2010 Analyzer or a commercial ELISA assay (Mercodia AB, Uppsala, Sweden). HbA1c was measured in EDTA whole blood using a Tosoh G7 Automated HPLC Analyzer, (Tosoh Bioscience, Inc, South San Francisco, CA 94080).

*JHS*

Jackson Heart Study (JHS) recruited 5306 African American residents living in the Jackson, Mississippi, metropolitan area of Hinds, Madison, and Rankin Counties. Participants were enrolled from 4 recruitment pools: random, 17%; volunteer, 30%; currently enrolled in the ARIC Study, 31% and secondary family members, 22%. Recruitment was limited to non-institutionalized adult African American men and women, 35-84 years old, except in a nested family cohort where those 21 to 34 years of age were also eligible. Among these participants, approximately 3400 gave consent that allows genetic research.

JHS participants received three back-to-back clinical examinations (Exam 1, 2000-2004; Exam 2, 2005-2008; and Exam 3, 2009-2013) that have generated extensive longitudinal data on traditional and putative cardiovascular disease risk factors and measures of subclinical cardiovascular disease. Exam 4 is ongoing. Biological samples (i.e., blood and urine) have been assayed for putative biochemical risk factors and stored for future research. DNA has been extracted and lymphocytes have been cryopreserved for genetic and omics studies.

Fasting (≥8 hours) plasma was used for glucose and insulin measurement, as described in PMID 15367870. Insulin was assayed at University of Minnesota using a radioimmunoassay kit from Linco; glucose was Assessed at the UMMC on a Vitros 950 or 250, Ortho-Clinical Diagnostics analyzer (Raritan, New Jersey). HbA1c was assessed using a high-performance liquid chromatography system (Tosoh Corporation, Tokyo, Japan) at University of Minnesota.

*MESA*

The Multi-Ethnic Study of Atherosclerosis (MESA) is a longitudinal cohort study of subclinical cardiovascular disease and risk factors that predict progression to clinically overt cardiovascular disease or progression of subclinical disease. Between 2000 and 2002, MESA recruited 6814 men and women 45–84 years of age from Forsyth County, North Carolina; New York City; Baltimore; St. Paul, Minnesota; Chicago; and Los Angeles. Participants at baseline were 38% White, 28% African-American, 22% Hispanic, and 12% Asian (primarily Chinese) ancestry, without evidence of clinical cardiovascular disease at baseline. Baseline data were collected using standardized questionnaires, physical examinations, and fasting laboratory blood tests.

At the baseline examination, participants completed a 120-item Block-style food-frequency questionnaire (FFQ) modified to include Chinese and Hispanic foods to accommodate the MESA population. The FFQ inquired about serving size (small, medium, large) and frequency of intake for selected foods and beverages (from “rare or never” to a maximum of “≥2 times/day” for foods and a maximum of “≥6 times/day” for beverages). The questionnaire also inquired on frequency, dosage, and duration of supplement use, allowing quantification of nutrient intake from supplements. Daily nutrient intake from foods was estimated by multiplying the reported amount consumed by its nutrient content (Nutrition Data Systems for Research; University of Minnesota; Minneapolis). All nutrients were adjusted for total energy intake using the residual method.

Participants fasted for 12 hours and avoided smoking and heavy physical activity for 2 hours before each examination. Fasting blood samples were drawn between 7:30 and 10:30 AM. Serum was frozen and stored at −70 °C. Information about socioeconomic status, medical history, medication, and tobacco and alcohol use was obtained through a questionnaire. Height and weight was measured by a stadiometer and calibrated scale. BMI was calculated from height and weight as kg/m^2^. Resting BP was measured three times, with participants in the seated position, with a Dinamap model Pro 100 automated oscillometric sphygmomanometer (Critikon); the average of the last two measurements was used in the analysis. Fasting TGs were measured in plasma after an 8-h fast using a glycerol-blanked enzymatic method (Trig/GB; Roche Diagnostics, Indianapolis, IN). HDL-C was measured in plasma by the cholesterol oxidase method (Roche Diagnostics) after precipitation of non–HDL-C magnesium/dextran. Serum glucose was measured using the glucose oxidase method on the Vitros analyzer (Johnson & Johnson, Rochester, NY). Diabetes status was defined according to the 2003 American Diabetes Association criteria of FG ≥ 7.0 mmol/l (126 mg/dl) or use of hypoglycemic medication (oral agents and/or insulin) as previously described(1,2).

*WHI*

WHI is a long-term, prospective, multi-center cohort study that investigates post-menopausal women’s health(3). WHI was funded by the National Institutes of Health and the National Heart, Lung, and Blood Institute to study strategies to prevent heart disease, breast cancer, colon cancer, and osteoporotic fractures in women 50-79 years of age. WHI involves 161,808 women recruited between 1993 and 1998 at 40 centers across the US. The study consists of two parts: the WHI Clinical Trial which was a randomized clinical trial of hormone therapy, dietary modification, and calcium/Vitamin D supplementation, and the WHI Observational Study, which focused on many of the inequities in women’s health research and provided practical information about the incidence, risk factors, and interventions related to heart disease, cancer, and osteoporotic fractures.  The participants for WHI who were included in TOPMed were selected for stroke, VTE, or as controls.

Dietary intake was assessed using a food frequency questionnaire (FFQ) designed specifically for WHI(4) to collect usual diet (reference period of prior three months). This FFQ included 122 line items for foods and food groups, 19 adjustment questions that add precision to estimates of fat intake and whole grain intake, and two summary questions. The FFQs were processed with the Nutrition Data Systems for Research (NDSR®, version 2005, University of Minnesota), which derives its data from the USDA Standard Reference database and is augmented with information from food manufacturers.

Glucose and insulin were measured in fasting (≥ 8 hours of fasting) serum or EDTA plasma samples.
