## Supplementary Tables for "Investigating gene-diet interactions impacting the association between macronutrient intake and glycemic traits"

**Supplementary Table S1:** Population characteristics, organized by cohort.

| **Cohort** | **N** | **Sex**  **(% female)** | **Age** | **BMI (kg / m^2^)** | **Population** | **Carbohydrate (%)** | **Fat (%)** | **FG (mmol/L)** | **FI (pmol/L)** | **HbA1c (%)** |
| --- | --- | --- | --- | --- | --- | --- | --- | --- | --- | --- |
| WHI | 6562 | 100% | 66.7 (6.9) | 28.1 (5.8) | Other: 22 (0%); American Indian: 35 (1%); Asian: 122 (2%); Hispanic/Latino: 232 (4%); African: 951 (14%); European: 5200 (79%) | 50.1 (9.1) | 33 (8.3) | 5.7 (0.6) | 54.3 (36.5) | 5.2 (0.4) |
| HCHS SOL | 6059 | 57.90% | 44.2 (13.8) | 29.4 (6.2) | Hispanic/Latino: 6059 (100%) | 51.7 (3.1) | 30.4 (2.6) | 5.2 (0.5) | 72.9 (53.4) | 5.5 (0.4) |
| ARIC | 5832 | 56% | 53.6 (5.7) | 26.7 (4.8) | African: 1102 (19%); European: 4730 (81%) | 48.9 (9.3) | 32.8 (6.6) | 5.4 (0.5) | 61.5 (44.8) | 5.4 (0.4) |
| MESA | 3621 | 51.60% | 60.8 (9.9) | 27.8 (5.1) | Hispanic/Latino: 766 (24%); African: 786 (25%); European: 1609 (51%) | 53.8 (8.7) | 30.4 (6.8) | 4.9 (0.6) | 37.7 (25.5) | 5.4 (0.4) |
| FHS | 2925 | 54.60% | 48.5 (11.9) | 26.9 (4.9) | Other: 2 (0%); European: 2923 (100%) | 49.4 (8.9) | 30.8 (6.5) | 5.2 (0.5) | 179.1 (89.4) | 5.5 (0.3) |
| CHS | 2693 | 59.20% | 76.6 (5.5) | 26.4 (4.4) | Hispanic/Latino: 27 (1%); African: 345 (13%); European: 2314 (86%) | 55.1 (8.2) | 29.9 (6.5) | 5.4 (0.6) | 98.2 (59.8) | N/A |
| CARDIA | 2156 | 56.80% | 45.2 (3.6) | 28.7 (6.3) | African: 880 (41%); European: 1276 (59%) | 47.3 (9.4) | 36 (8.1) | 5.2 (0.5) | 66.1 (38.9) | 5.3 (0.4) |
| JHS | 1331 | 62.50% | 52.9 (12.7) | 31.1 (7) | African: 1331 (100%) | 48.7 (9) | 36 (6.9) | 5 (0.5) | 94.9 (54) | 5.5 (0.5) |
| GOLDN | 807 | 53.30% | 46.5 (15.8) | 27.8 (5.4) | European: 807 (100%) | 49.2 (8.1) | 35.4 (6.4) | 5.4 (0.5) | 91.5 (50.2) | N/A |
| GeneSTAR | 426 | 63.40% | 45.8 (7.2) | 29.8 (6.2) | European: 162 (38%); African: 264 (62%) | 45.5 (8.5) | 38.3 (7.2) | 5 (0.7) | 57.9 (38.3) | N/A |

**Supplementary Table S2:** Study-specific regression modeling notes.

| **Study** | **Notes** |
| --- | --- |
| ARIC | No study-specific notes |
| CARDIA | No study-specific notes |
| CHS | Additional adjustment for study center and study center*CHO interactions; no HbA1c available |
| FHS | Additional adjustment for generation; no adjustment for population (single race/ethnicity) |
| GeneSTAR | No HbA1c available |
| GOLDN | No adjustment for population (single race/ethnicity); no HbA1c available |
| HCHS SOL | Additional adjustment for study center and study center*CHO interactions; no adjustment for population (single race/ethnicity); additional random effects for household and sampling block |
| JHS | No adjustment for population (single race/ethnicity) |
| MESA | No study-specific notes |
| WHI | No adjustment for sex (single sex); no random effect for kinship nor heterogeneous variances (no related individuals); no HbA1c available |

**Supplementary Table S3:** Single-variant GDI results passing a suggestive significance threshold of p < 1×10^-5^.

| **Trait** | **rsID** | **Chr.** | **Position** | **Non-Effect Allele** | **Effect Allele** | **Avg. EAF** | **Interaction estimate** | **P-interaction** | **P-joint** |
| --- | --- | --- | --- | --- | --- | --- | --- | --- | --- |
| HbA1c | rs79762542 | 4 | 77979164 | A | G | 0.03 | 0.0191  [0.0126-0.0257] | 1.14E-08 | 4.27E-08 |
| FG | rs1288694 | 3 | 71275429 | T | C | 0.61 | 0.00643  [0.00419-0.00868] | 1.93E-08 | 1.03E-07 |
| lnFI | rs782681704 | X | 155084576 | T | G | 0.01 | 0.113  [0.0728-0.154] | 4.57E-08 | 2.62E-07 |
| HbA1c | rs550200127 | 8 | 142474223 | A | G | 0.99 | 0.107  [0.0686-0.146] | 5.64E-08 | 1.50E-08 |
| HbA1c | rs146242010 | X | 153909957 | T | G | 0.96 | -0.0112  [-0.0155--0.00687] | 3.79E-07 | 1.05E-64 |
| HbA1c | rs142815083 | 7 | 148529092 | A | T | 0.02 | -0.0408  [-0.0575--0.0242] | 1.47E-06 | 5.21E-09 |
| HbA1c | rs624 | X | 154521720 | T | C | 0.1 | 0.0077  [0.00435-0.011] | 6.37E-06 | 8.37E-100 |

Interaction estimates with 95% CIs are given in units of [trait units / allele / 100 kcal carbohydrate]. P-joint corresponds to a joint, 2-degree of freedom test of the genetic main and interaction effects.

**Supplementary Table S4:** Gene-based aggregate rare variant testing results passing a suggestive significance threshold of p < 1×10^-4^.

| **Trait** | **Gene** | **Variant mask** | **# variants** | **# cohorts** | **Chromosome** | **P-interaction** | **P-joint** |
| --- | --- | --- | --- | --- | --- | --- | --- |
| FG | ENSG00000239620 | MS | 5 | 9 | 3 | 1.65E-05 | 1.54E-04 |
| HbA1c | ENSG00000150687 | MS | 6 | 6 | 11 | 2.76E-05 | 3.15E-04 |
| FG | ENSG00000101470 | RV | 2 | 9 | 20 | 3.93E-05 | 1.75E-04 |
| FG | ENSG00000265097 | RV | 2 | 2 | 18 | 4.18E-05 | 4.43E-05 |
| FG | ENSG00000204296 | MS | 7 | 10 | 6 | 4.83E-05 | 1.84E-04 |
| FG | ENSG00000105655 | RV | 5 | 10 | 19 | 5.22E-05 | 1.93E-04 |
| HbA1c | ENSG00000170476 | MS | 3 | 6 | 5 | 7.08E-05 | 2.87E-04 |
| HbA1c | ENSG00000157884 | MS | 2 | 6 | 2 | 8.85E-05 | 5.11E-04 |

P-joint corresponds to a joint, 2-degree of freedom test of the genetic main and interaction effects.

**Supplementary Table S5:** Results from the UKB replication analysis in the full unrelated sample.

| **rsID** | **Non-Effect Allele** | **Effect Allele** | **Discovery trait** | **Discovery GxCHO sign** | **UKB trait** | **Interaction estimate** | **P-value** | **EAF** |
| --- | --- | --- | --- | --- | --- | --- | --- | --- |
| rs79762542 | A | G | HbA1c | + | HbA1c | 0.29 | 0.025 | 0.0015 |
| rs79762542 | A | G | HbA1c | + | FG | 0.15 | 0.013 | 0.0055 |
| rs1288694 | T | C | FG | + | HbA1c | 0.01 | 0.642 | 0.6119 |
| rs1288694 | T | C | FG | + | FG | 0 | 0.679 | 0.6123 |
| rs782681704 | T | G | lnFI | + | HbA1c | -0.11 | 0.758 | 3.00E-04 |
| rs782681704 | T | G | lnFI | + | FG | 0.13 | 0.877 | 4.00E-04 |
| rs550200127 | A | G | HbA1c | + | HbA1c | -0.06 | 0.889 | 0.9998 |
| rs550200127 | A | G | HbA1c | + | FG | 63.3 | 0.172 | 0.9997 |
| rs146242010 | T | G | HbA1c | - | HbA1c | -0.16 | 0.118 | 0.9978 |
| rs146242010 | T | G | HbA1c | - | FG | -0.08 | 0.067 | 0.9915 |
| rs142815083 | A | T | HbA1c | - | HbA1c | -0.39 | 0.124 | 5.00E-04 |
| rs142815083 | A | T | HbA1c | - | FG | 0.13 | 0.32 | 0.002 |
| rs624 | T | C | HbA1c | + | HbA1c | -0.01 | 0.782 | 0.0075 |
| rs624 | T | C | HbA1c | + | FG | 0.02 | 0.432 | 0.021 |

Interaction estimates are given in units of [trait units / allele / 100 kcal carbohydrate]. Units are mmol/mol for HbA1c and mmol/L for FG.

**Supplementary Table S6:** Results from the UKB replication analysis in the African-ancestry-specific sample.

| **rsID** | **Non-Effect Allele** | **Effect Allele** | **Discovery trait** | **Discovery GxCHO sign** | **UKB trait** | **Interaction estimate** | **P-value** | **EAF** |
| --- | --- | --- | --- | --- | --- | --- | --- | --- |
| rs79762542 | A | G | HbA1c | + | HbA1c | 0.23 | 0.287 | 0.1116 |
| rs79762542 | A | G | HbA1c | + | FG | 0.15 | 0.046 | 0.1047 |
| rs1288694 | T | C | FG | + | HbA1c | 0.02 | 0.886 | 0.7024 |
| rs1288694 | T | C | FG | + | FG | 0 | 0.938 | 0.7136 |
| rs782681704 | T | G | lnFI | + | HbA1c | -0.34 | 0.68 | 0.0083 |
| rs782681704 | T | G | lnFI | + | FG | -1.24 | 0.487 | 0.0046 |
| rs550200127 | A | G | HbA1c | + | HbA1c | 0.21 | 0.842 | 0.994 |
| rs550200127 | A | G | HbA1c | + | FG | 15.41 | 0.538 | 0.9942 |
| rs146242010 | T | G | HbA1c | - | HbA1c | -0.11 | 0.497 | 0.8119 |
| rs146242010 | T | G | HbA1c | - | FG | -0.15 | 0.009 | 0.8283 |
| rs142815083 | A | T | HbA1c | - | HbA1c | -0.68 | 0.096 | 0.035 |
| rs142815083 | A | T | HbA1c | - | FG | 0.22 | 0.186 | 0.0406 |
| rs624 | T | C | HbA1c | + | HbA1c | -0.03 | 0.755 | 0.3913 |
| rs624 | T | C | HbA1c | + | FG | 0.03 | 0.416 | 0.3743 |

Interaction estimates are given in units of [trait units / allele / 100 kcal carbohydrate]. Units are mmol/mol for HbA1c and mmol/L for FG.
